# A conversational artificial intelligence agent for medication reconciliation and review

**DOI:** 10.1101/2025.06.16.25329719

**Authors:** Rahul C Deo, Shinichi Goto, Tara Jain, Sarah Meier, Rahul Patel

## Abstract

Medication reconciliation, the process of creating an accurate medication list for a patient, is critical to patient safety and care quality but requires clinical expertise and time. Large language models (LLMs), with the ability to generate language and respond to diverse prompts, offer the potential to automate medication reconciliation and review, including via spoken conversation. We developed AMREC, the Atman Medication REconciliation Conversational AI agent. AMREC uses a fine-tuned version of Llama-3.1-8B-Instruct to standardize a patient’s medication list by extracting 18 elements from each prescription. A voice agent then follows two dialogue flows: 1) iterating through the medication list with identification and correction of discordances and 2) collecting and clarifying requisite details on any additional medications taken. The extraction model achieved an accuracy rate of 98.3% across prescription elements, and user testing demonstrated the conversational AI agent’s ability to confirm, correct, remove, and add new medications to a candidate list. With additional development, AMREC could be deployed in the context of frequent medication reconciliation, thereby improving patient care outcomes and reducing the high-cost burden of medication errors on the healthcare system.

## Introduction

Medication reconciliation, a process in which a patient’s current medication list is compared to a prior authoritative version, has been a long-standing priority for healthcare safety and quality by multiple agencies, including The Joint Commission and The Institute for Healthcare Improvement. ^1–3^ Process improvements have focused on resolving medication discrepancies at transitions of care to reduce adverse events that may occur when, upon discharge, essential medications the patient took at home before hospitalization are not restarted or new medications are not initiated.^4^ Although these transitions are a major source of potential error, many have emphasized that a more frequent and thorough medication review process is needed to identify discordances between prescribed and actual medication use and uncover underlying reasons such as side effects, costs, and lack of education, to avoid medication errors and improve patient adherence.^5–7^

Unsurprisingly, frequent medication review plays a critical role in systematized, software-driven medication management workflows, which have been adopted by technology-forward healthcare companies and population health programs.^8,9^ Our own company, Atman Health, has implemented a software-driven approach to the medical management of multiple conditions across primary and specialty care. ^10^ Given that an accurate medication regimen and historic side effects are essential inputs to our software logic, we have emphasized medication review at the onset and duration of our programs. We have found that confusion around medication lists occurs frequently, even outside of transitions of care, driven by multiple providers making medication changes (including primary care and specialists) without a centralized source of truth, and pharmacies sometimes dispensing outdated prescriptions, often through automated refill programs. Thus, active management of medications in the outpatient setting needs frequent medication review to be safe and successful.

Conducting an effective medication reconciliation can take 15-30 minutes or longer for elderly patients with more complex medication regimens.^11^ The need for an efficient and effective process has only increased as our global population rapidly ages and the number of new approved medications continues to rise. However, the time and expertise required for such a process, along with the resulting cost, present a challenge with no obvious solution.

Healthcare has begun to adopt generative artificial intelligence (AI)-based solutions to improve patient outcomes and reduce costs across a range of applications, including drug discovery, medical diagnosis, patient education, personalized medicine, healthcare administration, and medical education.^12^ Ambient scribes supporting clinical documentation are poised to become one of the fastest technology adoptions in the history of healthcare.^13^ The role of conversational AI (chatbots) in health care to date has been primarily focused on 1) delivery of remote services with patient support, care management, education/skills building, and health behavior promotion and 2) provision of administrative assistance to health care providers.^14–16^ Some companies are venturing into the space of patient-facing conversational AI agents. However, these agents are primarily adopting roles removed from the process of active medication changes^17^ and, as such, may have limited utility in improving clinical outcomes. ^18^

We sought to develop a conversational AI agent to facilitate the medication review process used in active medication management. The objective would be to create an agent capable of creating, reviewing, and updating medication lists, as well as discussing barriers to adherence. For efficacy, the agent would need to be able to work with the full complexity of existing prescription signatures as well as the range of a complete compendium of medications.

## Methods

### Ethics

The project was conducted as a quality improvement initiative aimed at enhancing the consistency and quality of our company’s medication reconciliation process.

### Structured representation of prescription data

Prescription orders consist of a prescription unit (medication name, strength, dose form – e.g., lisinopril 40 mg tablet) and a "signature", which includes instructions on how it should be taken (e.g., 1 tablet daily in the morning). Although the prescription unit is standardized, mapping to a namespace such as RxNorm or NDC, the signature is not. Given that a conversational agent (and any downstream computed logic) requires a structured representation of data to determine what is missing, our first task was to model the prescription signature.

We reviewed prior publications ^19,20^, as well as 970 deidentified prescription signatures collected through this QI improvement effort. We identified 18 distinct elements to capture within a prescription (Table 1). The simplest prescriptions might only have three of these, while more complex prescriptions might have 10 or more.

**Table 1:** Elements of prescription.

| Element | Description | Example |
| --- | --- | --- |
| no_taken | Number of units taken | Take **1** tablet once daily |
| no_taken_1 | If 2 or more different number of units are taken, the first value | Inject **4** units with breakfast, 5 units with lunch, and 9 units with dinner |
| no_taken_2 | If 2 or more different number of units are taken, the second value | Inject 4 units with breakfast, **5** units with lunch, and 9 units with dinner |
| no_taken_3 | If 3 or more different number of units are taken, the third value | Inject 4 units with breakfast, 5 units with lunch, and **9** units with dinner |
| no_taken_ll | If a range of number of units are taken, the value of the lower limit | Take **1-2** capsules as needed for pain. |
| no_taken_ul | If a range of number of units are taken, the value of the upper limit | Take 1-**2** capsules as needed for pain. |
| no_taken_dur_2 | If two different time durations are specified, the number taken for the second duration | Take 1 tablet three times a day for 5 days and then **1** tablet daily at bedtime for 90 days. |
| freq | The frequency at which the medication is taken | Take 1 tablet **once daily** |
| freq_1 | If 2 or more different number of units are taken, the second frequency – typically ellipted, defaults to once a day | Take 1 tablet (**...once daily...**) in the morning and 2 tablets in the evening. |
| freq_2 | If 3 or more different number of units are taken, the second frequency – typically ellipted, defaults to once a day | Take 1 tablet in the morning and 2 tablets (**...once daily...**) in the evening. |
| freq_dur_2 | If two different time durations are specified, the frequency the medication is taken for the second duration | Take 1 tablet three times a day for 5 days and then 1 tablet **once daily** for 90 days. |
| time_of_day | The time of day the medication is taken | Inject 4 units **with breakfast**, 5 units with lunch, and 9 units with dinner |
| time_of_day_2 | If the medication is taken at two or more times of the day, the second time of day | Inject 4 units with breakfast, 5 units **with lunch**, and 9 units with dinner |
| time_of_day_3 | If the medication is taken at three or more times of the day, the third time of day | Inject 4 units with breakfast, 5 units with lunch, and 9 units **with dinner** |
| dur_1 | If two different time durations are specified, the number of days of the first duration | Take 1 tablet three times a day for **5 days** and then 1 tablet once daily for 90 days. |
| dur_2 | If two different time durations are specified, the number of days of the second duration | Take 1 tablet three times a day for 5 days and then 1 tablet once daily for **90 days**. |
| unit_taken | The unit taken | Take 1 **tablet** once daily |
| PRN | If the medication is taken as needed (boolean) | Take 1-2 capsules **as needed** for pain. |

### Design of a conversational AI agent

#### Clinical workflow

To build a conversational AI agent for medication reconciliation, we first needed to define the intended clinical workflow, focusing initially on our own business needs. Our workflow for a new patient involves following a series of steps to ensure that the medication list is a complete and structured representation of all intended prescriptions. Furthermore, a taking status is captured for each prescription:

- The medication is not taken

◦ The patient believes the provider discontinued the medication
◦ The patient is unaware that the provider prescribed the medication
◦ The patient thinks they should be taking the medication, but it was not refilled
◦ The patient elects not to take it due to side effects, cost, lack of perceived need, or other reasons
- The medication is taken differently than how it is prescribed
- The medication is taken as prescribed

The medication list is addended with prescription details of any additional medications taken by the patient.

The actual workflow to achieve this is currently carried out by human agents (Figure 1). A candidate medication list is imported from various sources, including data exchanges such as Surescripts, a pharmacy benefit manager, and Continuity of Care Documents (CCDs), which are electronic documents summarizing a patient’s health record. Whereas all prescription entries include a prescription unit, some (∼25%) are missing a signature. As described above, prescription signatures are in plain text form and have no standardized format.

**Figure 1.**
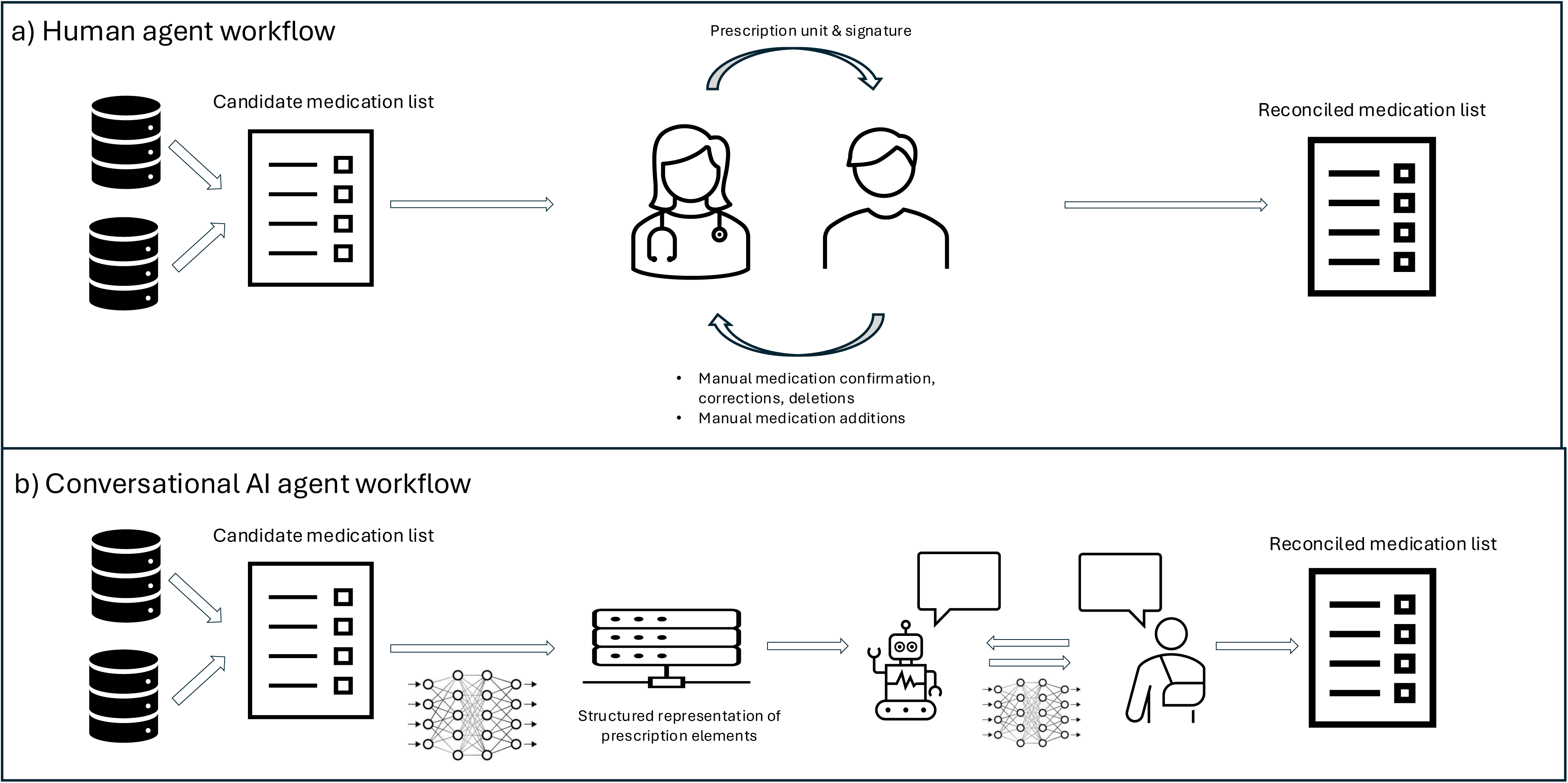
**Figure 1a.** The medication reconciliation workflow carried out by a human agent includes import from data exchanges to create a candidate medication list followed by 1) review of prescription unit and signatures on the candidate list with the patient, resulting in medication confirmation, corrections and deletions and 2) addition of patient-reported medications not on the candidate list, resulting in a reconciled medication list **Figure 1b.** The medication reconciliation workflow carried out by conversational AI agent also includes import from data exchanges to create a candidate medication list, followed by LLM extraction of prescription elements from unstructured signatures and AI-patient dialogue to iterate through the medication list and add new medications, resulting in a reconciled medication list.

In an encounter with the patient, the agent performs the following:

#### Review of medications on the candidate list

The agent will first have the patient confirm the prescription unit and then the prescription signature. The patient supplies any missing instructions. If the medication is taken differently, both the original instructions and the actual method of administration are captured. Finally, if the prescription unit has been changed (e.g., increase or decrease in strength), a new medication entry is created

#### Addition of patient-reported medications not on the candidate list

The patient is then asked if they are taking any additional medications. A new record is created for every additional proposed medication. To do so, the prescription unit is searched against a database, and confirmation is obtained for the name, strength, and dose formulation. Often, the patient will simply provide the medication name, and the agent then offers possible strengths to choose from. A failure to match a strength or dose formulation exactly may indicate a problem with the medication name. Combination medications, which include two or more ingredients (e.g., olmesartan/hydrochlorothiazide 20-12.5 mg tablet), can be misleading as the patient may only read the first ingredient in the name. These can sometimes be caught when the strengths do not match, though there remains a risk that the patient only matches one of the ingredient strengths. The directions on how to take the medication are provided. The agent has a list of standard instructions for most medications. If the proposed method of taking differs from the candidates, the patient is asked to clarify the situation.

The output of this process is an updated medication list consisting of an annotated version of the original list, as well as details on any additional medications.

### Replicating the process with a conversational AI agent

Task-oriented dialogue systems, colloquially termed "chatbots" have evolved rapidly in recent years due to marked improvement in large language models. Most frequently, they have been used in customer service applications ^21^. Historically, the approach has been to couple three components: a natural language understanding module, which takes a user’s statement and classifies it to one of several predefined intents, a dialogue management module, which contains the business logic to gathering the requisite details to address the intent, and a natural language generation module, which produces speech ^22^. This approach, although powerful, suffers foremost from the challenge of classifying intents, which can be ambiguous.

With the development of large language models, some have hoped that the entire process can be delegated to an oV-the-shelf language model agent, which has no pre-specified business logic and just engages in a free-form conversation with the user. Although this approach may work for generic activities, such as looking up details, it is unlikely to perform well in specialty applications where the required data and potential dialogue flows differ substantially from the training data the model has seen.

We sought a hybrid approach that enables the collection of pre-specified data (i.e., prescription information) and follows the successful flows we have designed but with the flexibility to accommodate inevitable deviations that arise in natural dialogue. The main deviations we experience are:

1. Corrections: the user realizes the prior statement was incorrect and proposes a correction. In the conversational Analysis field, this is termed "conversation repair" ^23^ .
2. Digression: the conversation sparks a digression, such as a discussion of a previous hospital admission.
3. Mismatching: the user states a medication name that is either incorrectly spelled (if spelled), misspelled by the speech-to-text model, or does not match one of the medication reference names in the database exactly.

After reviewing potential frameworks, we selected Rasa^24^, an open-source Python-based framework that prespecifies business logic and dialogue flows for the intended use or "happy paths," and uses large language model-driven conversation to address deviations ^24^ . We termed our application "AMREC" for Atman Medication REconciliation Conversational agent (Figure 1b).

### An LLM-focused approach to medication reconciliation

The medication reconciliation process, in concept, is simple. A complete prescription consists of a series of data elements. For known prescriptions, the conversation with the patient begins by ensuring alignment on the prescription unit being considered (by matching the values of the prescription unit data elements) and then confirming that they are following the prescription instructions (by again matching the values of the prescription units). If discrepancies occur, including not taking the medication at all, the specific data elements are updated. Finally, a status is captured regarding whether they are taking the medication infrequently. For a conversational AI agent to achieve this, it must be able to propose various elements or combinations of elements to the patient and understand the dialogue sufficiently to extract agreement or disagreement and the values of the relevant data elements.

For patient-reported prescriptions, the AI agent must be able to solicit the data elements required for a complete prescription. Depending on the complexity of the medication (e.g., multiple ingredients or not) and the complexity of the signature (e.g., different number of units taken at different times, one or more defined durations), this may require a different series of questions.

Rasa’s approach to this problem is to use LLM models to 1) recognize which dialogue flow is desired by the user, 2) perform the data element extraction from natural dialogue, and 3) recognize unhappy paths in the conversation that require some additional activity such as chit-chat before returning to the desired data or seeking help from a human agent. To achieve this, a structured prompt is fed to the LLM that contains a description of the dialogue flows, the data elements including permitted fields, the conversation that occurred to date, the current values of data elements, the last proposed question, the last user answer, as well as general guidance on what the LLM is asked to produce as output. The output of the LLM is a series of structured actions. For example, if the desired action is to set a value to a slot for the strength of the medication, the output would be "SetSlot(strength, 5)".

### Defining data elements

We started with the 18 prescription elements described above. In addition, we added the following data elements per prescription:

1. Prescription category: whether the prescription was listed or patient-reported
2. Confirmation status: prescription unit, prescription signature, strength, dose formulation, number taken, and frequency
3. Reason if not taking the medication: provider discontinued or didn’t refill it; patient decided not to take it
4. Adherence status: yes or no, if missing doses

### Defining dialogue flows

The primary flows are as follows (Figure 2):

**Figure 2:**
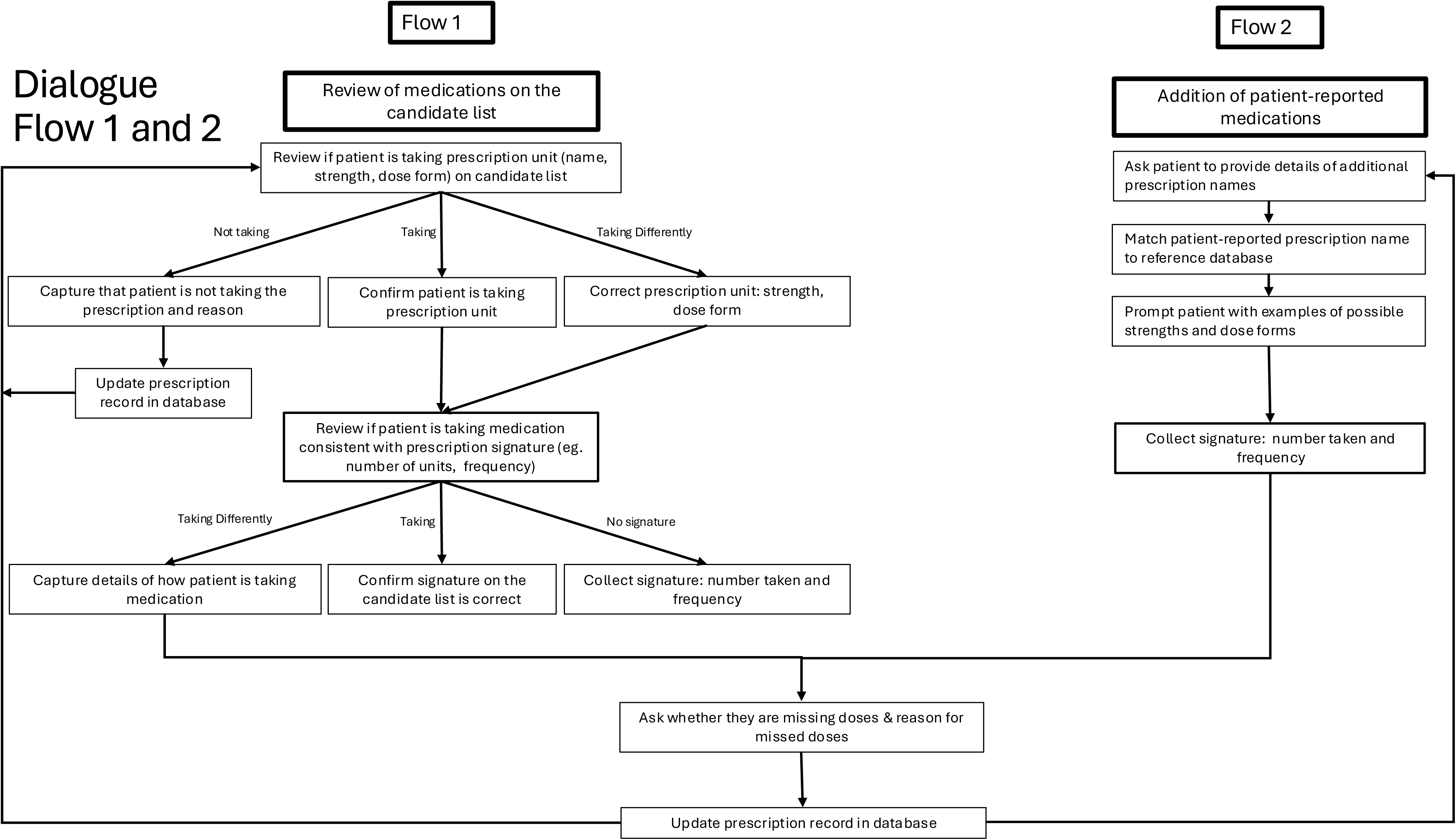
Dialogue Flows 1 and 2 with the goal of confirming a list of medications with an accurate prescription unit (e.g. name, strength, dose form), prescription signature (e.g. number of units, frequency), and details on patient adherence.

Flow 1: Review of medications on the candidate list

Flow 2: Addition of patient-reported medications

#### Flow 1: Review of medications on the candidate list

For each entry in the medication list, the AI agent closely follows the clinical flow defined earlier. The user is asked to confirm the prescription unit, followed by the prescription signature. If there is any discrepancy in the prescription signature, either regarding the number of units taken or the frequency, they are asked to provide a correction. If there is any discordance in the medication strength or dose formulation, a new medication entry is created, and Flow 2 is followed.

If a prescription signature is missing, the user is asked to enter the signature *de novo*, which resembles Flow 2. A critical branching point occurs when a different number of units is taken at one time of day compared to another (for example, 1 tablet in the morning and 2 tablets at night). The user is also asked if the medication is taken as needed.

If edits are made to the prescription signature, a confirmation of the signature is performed, and then a final confirmation of the entire prescription. Once confirmed, a question is asked regarding any missed doses (to infer adherence challenges). Finally, the AI agent updates the prescription record in the database and moves to the next prescription in the list.

Function calls needed for flow 1 include:

1. Selecting an entry from the medication list
2. Updating the database record of the prescription
3. Converting the prescription data fields into a sentence to repeat back to the patient.

#### Flow 2: Addition of patient-reported medications

The flow is inherently more challenging because the patient must provide complete details of the prescription, rather than simply confirming the details presented to them. If they are reading from a prescription bottle, there may be abbreviations of the medication name and the dose formulation, and either a brand or generic name may be used. To our knowledge, prescription names on either printed medication lists (supplied by the electronic medical record) or prescription bottles do not follow any standardized namespace.

The patient may also inadvertently omit some details of the prescription, believing them to be unimportant. If this flow occurs via typed chat, there is the risk of misspelling. If it is performed via telephone, there is an additional risk of mispronunciation.

The flow we use here is very similar to the human agent flow described above. The patient is asked to first focus on the medication name only, though often they may also provide the strength, unit of strength, and dose formulation. The AI agent extracts the medication string and searches against a database for candidates. If no perfect match is identified, the user is presented with medication name candidates. If they select one with an identical match, candidate strengths are then provided for them to choose from. If there is no match, they are again asked to re-supply the medication name. If they choose not to or if an additional strength matching cycle fails, the user is then asked to proceed to the following medication. If a medication name and strength match is found, they are asked to confirm the dose formulation. If all three match, the prescription unit is validated, and the user is asked to provide the prescription signature and whether the medication is taken as needed. Once the prescription is confirmed, the patient is asked about any missed doses.

Function calls needed for flow 2 include:

1. Medication string matching against a database

a. Extraction of candidate medication names
b. Extraction of candidate medication strengths
c. Extraction of candidate dose formulations.
2. Updating the database record of the prescription
3. Converting the prescription data fields into a sentence to repeat back to the patient.

### Software tests

We started with a series of 179 tests to cover the entirety of both flows. A test consists of a dialogue between a user and an AI agent where the expected behavior of the agent in response to the user’s last utterance is evaluated. The agent follows the business logic defined by the flows, while the user can respond in brief points (e.g., lisinopril), phrases, or complete sentences (e.g., "The medication I take is lisinopril. I absolutely love it."). Tests evaluate whether the correct element was captured from the dialogue and whether the following software-proposed action was correct.

Tests involved introducing or varying the following:

1. Prescription unit details: medication names, strengths, dose formulations
2. Prescription signature details: number of units taken, frequency, duration, PRN
3. User wording in describing these items, including variation between complete sentences (as expected in a spoken dialogue) and brief bullet entries (as expected in a typed chat)
4. AI agent wording in requesting the data
5. Variation in medication complexity: e.g., double and triple ingredient medications such as olmesartan-amlodipine-hydrochlorothiazide
6. Variation in prescription signature complexity: e.g., 3 different numbers of units taken at 3 different times of the day for a fixed number of days.
7. Misspellings of medication names
8. User digressions
9. User answering with information on multiple requested items despite being asked for a single value (e.g., number taken and frequency given, when only number taken is requested).

Testing was designed to highlight areas of deficiency in the model, thereby focusing fine-tuning.

### Fine-tuning

Although oV-the-shelf LLMs are effective at understanding dialogue, they may not be optimized for more specialized tasks, such as working with sentences that contain medication names and other prescription details. To improve the performance of our model, we fine-tuned it using a variant of Meta’s Llama-3.1-8B-Instruct as the base model (https://huggingface.co/rasa/cmd_gen_llama_3.1_8b_calm_demo). Training data, consisting of prompts and completions, were based on the original tests, with augmentation through variations in user responses, AI agent phrasing, and prescription details.

The model was trained iteratively using mlx-lm (https://ml-explore.github.io/mlx/) and LORA^25^ for 2750 iterations on a Mac Studio with 64GB RAM and an M1 Max processor, with a batch size of 16, a learning rate of 1x10-5, 16 LORA layers, a rank of 8, alpha of 16, scale of 10, and gradient- checkpointing. An independent test set was used for evaluation.

### An LLM to standardize imported prescriptions

As is clear from the data element requirements of AMREC, the prescription signature must be in a structured form. To achieve this goal, we trained an LLM model that could extract the requisite inputs from unstructured prescription signatures. Our goal was to extract the same 18 individual elements described above (Table 1).

#### Simulated prescription signatures

For training data, we first inspected 965 deidentified prescription signatures to derive a general syntax. We then simulated 57,000 prescription signatures that contained subsets of these elements. We broadened the complexity by introducing an optional verb at the beginning of the sentence (e.g., take, inject) and an optional route (e.g., by mouth, orally, to the skin). For elements with numbers, we included examples that included decimals and integers and expressed numbers as either words or digits. Prescription signatures varied in complexity and could include variable numbers of units at different times of the day (e.g., 2 tablets in the morning and 1.5 tablets in the evening) and fixed durations (take 1 capsule three times a day for 14 days, followed by 1 capsule daily for 90 days).

#### An entity recognition model for prescription signature elements

We started with the fine-tuned Llama-3.1-8B-instruct model, which was trained for entity extraction in AMREC. We modified the prompt to focus on prescription signatures, aiming to capture a combination of 18 elements in a standardized format. For categorical variables such as frequency and time of day, we also required the model to map the element to an available choice. For example, the model was expected to capture a frequency of "once a day", even if the signature was written as "once daily" or "daily".

The model was trained iteratively using mlx-lm and LORA ^25^ for 1000 iterations on a Mac Studio with 64GB RAM and an M1 Max processor, with a batch size of 16, a learning rate of 1x10-5, 16 LORA layers, a rank of 8, alpha of 16, scale of 10, and gradient-checkpointing. An independent test set was used for evaluation.

### User testing

A total of two users evaluated AMREC. Each evaluation consisted of confirming or correcting 10 pre-established prescription signatures of varying complexity, as well as reporting two additional medications. Users were asked to perform a mixture of confirmation, non-confirmation, and data element correction flows.

#### User testing with a text conversational agent

Users were asked to interact via typing responses, which naturally resulted in briefer entries for AMREC to parse and may involve typos.

#### User testing with a speech conversational agent

Text-based interaction, although valuable, fails to reflect the additional complexity of spoken dialogue. These include longer user utterances, potential mispronunciations – especially of medical terms – and a greater tendency towards digression. To test AMREC in the context of actual conversation, we incorporated an API to Deepgram and Cartesia’s voice-to-text, text-to-voice services into AMREC.

## Results

### A trained LLM can standardize prescription signatures

The fine-tuned Llama-3.1-8B-instruct model achieved a total accuracy of 98.3% across the 18 elements. Some variation was observed in performance, with slightly weaker results (95-97%) for elements in the second time period when the prescription signature included two distinct durations (e.g., freq_dur_2, no_taken_dur_2).

### A conversational AI agent – AMREC - can reconcile medications through dialogue

The conversational AI agent, AMREC, was developed to facilitate two dialogue flows: confirmation of existing medications and patient reports of additional medications. A series of 179 tests were designed to determine whether AMREC could accurately capture the requisite prescription elements from patient dialogue. Using the base model (rasa/cmd_gen_llama_3.1_8b_calm_demo), 75 of 179 failed. Failures were widespread but predominantly centered on the challenges of capturing natural patterns of speech, where a single sentence may include multiple elements, which conflicts with the original Rasa model design that emphasizes capturing elements one by one. Mapping elements to categorical variables (e.g., frequency, time of day) also had a high failure rate.

Fine-tuning via LORA was able to overcome deficiencies in the original model. The fine-tuned Llama-3.1-8B-instruct model failed 5 of 179 tests, with failures related to broken conversation patterns, predominantly user digressions.

### User testing confirms the eDicacy of the AI agent

Two users tested the software using various scenarios. Representative screenshots of dialogue in the chat format are shown (Figure 3). These include examples where prescriptions are confirmed, medication strengths are corrected, users describe stopping medications, and users report new prescriptions with some parenthetical statements.

**Figure 3:**
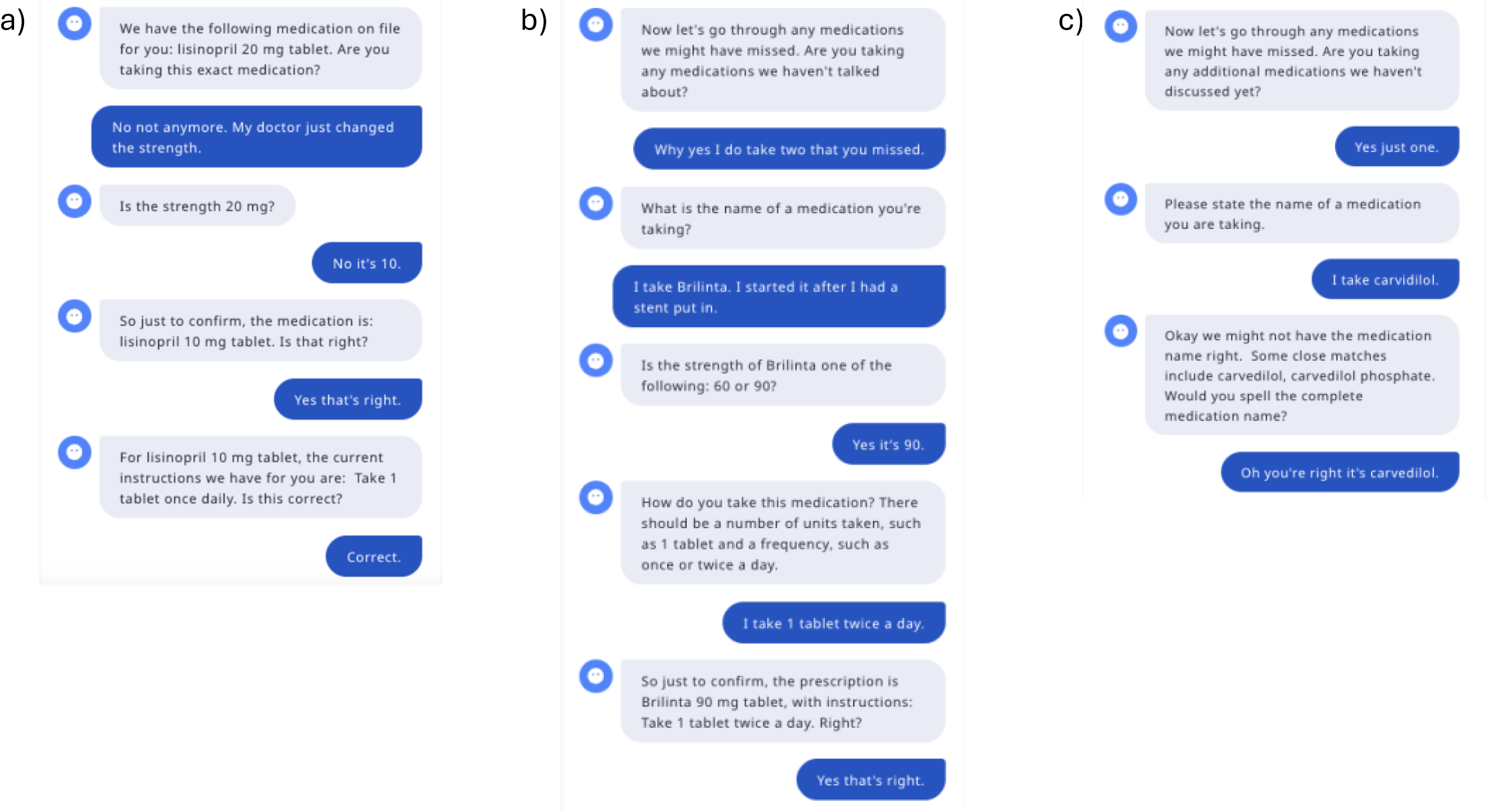
Screenshots of dialogue between AMREC and a patient user in chat format in which the a) user takes a different medication strength, and the prescription is corrected b) user reports a new prescription, and the prescription is added c) user misspelled the medication name and AMREC proposes alternatives

We also tested the model using speech rather than written chat. The primary challenge we identified was related to misspelled medication names, for which AMREC had to propose alternatives (e.g., Figure 3c). A recorded interaction between AMREC and a user, covering three listed prescriptions and one reported one, can be found here https://tinyurl.com/yb2xsze8.

## Discussion

We developed AMREC, a conversational AI agent that reconciles a medication list through dialogue, combining a structured dialogue framework with a trained LLM to extract prescription data elements and handle deviations in conversation. This Llama-3.1-8B-instruct model fine-tuned via LORA achieved 98.3% accuracy across the 18 prescription signature elements. Failures occurred in 5 of 179 tests due to deviations from expected dialogue patterns. User testing, including both text and speech inputs, confirmed AMREC’s ability to verify, correct, and add medications to the candidate list, even when users provided mispronounced or misspelled input.

Medication errors represent a leading cause of patient harm worldwide, associated with an annual cost of $42 billion, and can occur at all points in the care continuum—not limited to transitions of care.^26^ The review and reconciliation process has been demonstrated to reduce errors of omission, duplication, dosing errors, and drug interactions, but it remains a time- and people-intensive process for healthcare providers. LLMs have shown promise in healthcare applications with the ability to process complex concepts and respond to diverse prompts.^12,27–29^ A conversational AI agent for medication reconciliation may offer a compelling value proposition for healthcare providers, as it enables the automation of the medication reconciliation process and reduces costs. For our own Atman Health platform, which enables rapid iterative management of chronic diseases, frequent medication reconciliation is central to patient care to maximize patient adherence and avoid drug-drug interactions.^10^ AMREC can support our medication titration programs with increased operational efficiency at low cost.

One can evaluate the risk of a product such as AMREC by following the principles of the international standard on risk management in medical devices^30^ enumerating hazardous situations, hazards, and harms. The primary hazardous situation in any medication reconciliation process, especially one performed by a non-expert, is a false negative – a patient is taking a medication, but the process fails to capture it, either because of problems with name matching, the patient forgets to mention it, or there is some lack of comprehension. Prescribing in that setting increases the risk of a hazard from an undetected medication-medication interaction, such as duplication of a medication class (increasing risk of side effects) or in rarer cases, altering pharmacokinetics of an agent with a narrow therapeutic index. Efforts to move this solution into production will require extensive testing to estimate the rate of false positives and develop mitigation strategies against this occurrence.

This work has focused primarily on designing a standardized logical flow for medication reconciliation that can handle a full spectrum of complex prescription signatures. Further work is needed in three areas before deployment can occur. The first area is developing robustness to a greater diversity of number of unhappy paths likely to occur in conversation. Our experience as care providers has created an extensive compendium of such examples and additional cycles of fine-tuning should enable a greater robustness of the model. The second area involves implementing the model in an engineering stack that can demonstrate low latency of responses and handle broken conversations, including interruptions. Encouragingly, this has become an increasingly mature area with many vendors offering products to streamline this process. ^31^ Finally, medication names present an additional challenge, which includes the low likelihood that existing voice to text models can recognize these terms, and patients’ difficulties with pronunciation. Such models will need fine-tuning to reduce error rates. This flow should encourage spelling as well as a robust search against existing medication names to narrow the list of candidates. Refinement of this process aligns closely with the Atman Health platform, which is designed to support frequent and efficient medication reconciliation as a foundation for safe and scalable medication titration programs in chronic disease management.

## Data Availability

All data produced in the present study are available upon reasonable request to the authors

